# The association between Dietary Inflammatory Index and Latent Tuberculosis Infection in US participants:a cross-sectional study from NHANES

**DOI:** 10.1101/2025.06.19.25329974

**Authors:** Jingbo Jia, Yuanyuan Liu, Hua Zhang, Jianrui Pi, Chao Wang, Shiqi Dong, Tong Zhang, Wanjie Yang

## Abstract

**Background:** There is a growing recognition that inflammation related to diet could influence Latent Tuberculosis Infection(LTBI). However, the association between Dietary Inflammatory Index(DII) and LTBI remains unclear.

**Objective:** This study aimed to explore the relationship between DII and LTBI using a cross-sectional design, incorporating univariate and multivariate logistic regression analyses.

**Methods:** We conducted a cross-sectional analysis involving 3,892 participants from the National Health and Nutrition Examination Survey (NHANES).

Demographic data, including age, sex, race, body mass index (BMI), education level, poverty income ratio (PIR), marital status, smoking, alcohol consumption, hypertension (HPT), and diabetes mellitus (DM), were collected from all participants. We used logistic regression, smooth curve fitting, and subgroup analys to achieve our research objectives.

**Results:** Among all eligible participants, the mean age was 48.4 ± 17.8 years, with males comprising 50.6% of the study population. After adjusting for confounding variables, we found a positive association between the DII and the risk of LTBI, with an odds ratio of 1.07 (95% CI, 1.01–1.13; P=0.023). Adjusted smoothed plots also suggest a straightforward linear relationship between DII and LTBI(P for non-linearity = 0.924),as the level of DII increases, the risk of LTBI shows an upward trend.Additionally, subgroup analysis within the defined gender, age, BMI, PIR, smoking, drinking, HPT, and DM groups indicated no significant interactions among the subgroups, as evidenced by all P values for interaction exceeding 0.05.

**Conclusions:** These results underscore that DII may serve as an important risk factor for LTBI, providing new insights for early screening and intervention strategies. However,further studies are needed to explore the underlying mechanisms.

## Introduction

The worldwide impact of tuberculosis (TB) continues to pose a considerable public health issue[1–3], with Latent Tuberculosis Infection(LTBI) serving as a critical reservoir for potential future active TB cases[4–5]. LTBI is characterized by the presence of mycobacterium tuberculosis in individuals who do not show any clinical symptoms and are not capable of transmitting the infection to others.

According to the World Health Organization (WHO), it is estimated that around one-quarter of the global population is infected with LTBI, which significantly increases the risk of reactivation into active TB disease, especially among at-risk groups[6]. This unrecognized epidemic highlights the urgent need for effective predictors that enable early identification to prevent progression from LTBI to active TB to reducing the associated healthcare burden[7–9].

Recent studies have begun to explore the relationship between diet, inflammation, and the immune response. Diet and individual nutrients can influence systemic markers of immune function and inflammation[10–12], and may also influence people’s TB related health outcomes.A growing body of evidence suggests that diet can significantly affect immune responses, potentially altering the course of chronic diseases. For instance, diets high in processed foods and sugars are associated with increased systemic inflammation, which may impair the immune system’s ability to combat chronic diseases [13]. Conversely, diets rich in fruits, vegetables, and whole grains have been associated with lower levels of inflammatory markers, suggesting a protective effect against chronic inflammatory conditions and possibly enhancing immune function [14].

The DII is a tool developed to assess the inflammatory potential of an individual’s diet. However, the association between DII and LTBI remains unclear. Thus, the aim of this study was to explore the clinical and predictive significance of DII and LTBI in US people.

## Materials and Methods

This cross-sectional study employed data from the NHANES for the years 2011 to 2012, which was administered by the Centers for Disease Control and Prevention [15]. The primary objective of the NHANES program was to evaluate the health and nutritional status of non-institutionalized individuals residing in the United States through a structured multistage probability sampling approach [16]. The NHANES collects extensive demographic and health-related information utilizing home visits, health screenings, and laboratory tests conducted at a mobile examination center (MEC). The study received ethical approval from the Ethics Review Committee of the National Center for Health Statistics (NCHS), with all participants providing written informed consent prior to their participation. The secondary analysis of the data did not require additional Institutional Review Board approval [17]. The NHANES data can be accessed via the NHANES website (http://www.cdc.gov/nchs/nhanes.htm) (accessed on November 26, 2024). Our study included individuals who had undergone tuberculosis testing, while those with incomplete information regarding tuberculosis infection, DII, or other covariates were excluded from the analysis.

We conducted a characterization of LTBI within the NHANES datasets from 2011 to 2012 by employing two distinct evaluation methods: the tuberculin skin test (TST) and the interferon-gamma release assay (IGRA). A TST was classified as positive if the measurement of induration exceeded 10mm, accompanied by vesiculation and ulceration of the tuberculin purified protein derivative. The QuantiFERON® TB Gold IT assay serves as a diagnostic instrument for assessing cell-mediated immune responses to peptide antigens derived from mycobacterium proteins, which triggers the synthesis and release of the cytokine interferon-gamma (IFN γ). According to established criteria, the QuantiFERON® TB Gold assay is considered reactive if the nil value is ≤ 8.0 IU/ml, the TB antigen minus nil value is ≥ 0.35 IU/mL, and the TB antigen value minus nil value constitutes at least 25% of the nil value [17].Notably, regardless of the positivity of TST or IGRA, both tests were interpreted as indicators of LTBI [18].

The DII was first introduced by Shivappa et al. in 2014[19]. The index is calculated based on an individual’s dietary intake.Trained staff at the Mobile Inspection Center (MEC) conducted dietary recall interviews with participants, who were required to recall dietary intake data in the 24 hours.The DII for this study was calculated using 28 nutrients, including carbohydrates, proteins, fats, vitamins, and minerals.DII scores were calculated by summing all food component-specific DII scores.Higher DII scores indicate a greater dietary inflammatory potential, which is reflected in higher plasma levels of inflammatory biomarkers [20, 21], further studies confirmed with the increasing of circulating inflammatory biomarkers, including IL-1β, IL-6, TNF-α, and CRP, as well as elevated white blood cell (WBC) counts[22–24]. While a negative score indicates an anti-inflammatory potential. A score of zero means there is no significant effect on inflammatory potential.

A range of potential covariates was evaluated in accordance with the literature [25–27],encompassed a variety of demographic and health-related variables, including gender, age, race, educational attainment, marital status, smoking behaviors, alcohol consumption, BMI, DM, HPT, and PIR. Race was classified into several categories: Mexican American, non-Hispanic White, non-Hispanic Black, other Hispanic or other racial groups. Educational status was divided into two groups: individuals with less than a high school education and those with a high school education or higher. Marital status was defined as either married (including those who are married or cohabiting) or non-married (which encompasses individuals who are widowed, divorced, separated, or never married). Consistent with definitions from previous studies, smoking behavior was categorized into current smokers, former smokers (defined as those who have smoked more than 100 cigarettes in their lifetime and subsequently quit), and never smokers (those who have smoked fewer than 100 cigarettes). Alcohol consumption was categorized as either “yes” (indicating that the individual had consumed at least 12 alcoholic beverages within a single year) or “no.” The diagnoses of DM and HPT were determined based on participants’ responses to a questionnaire that inquired whether a medical professional had ever diagnosed them with these conditions. BMI was computed using a standardized method that considers both weight and height measurements.

This constitutes a secondary analysis of publicly available datasets. Categorical variables were expressed as proportions (%) while continuous variables were characterized by the mean (standard deviation, SD) or median (interquartile range, IQR), as suitable. To assess the disparities among groups, one-way analyses of variance (normal distribution), Kruskal–Wallis tests (skewed distribution), and chi-square tests (categorical variables) were conducted. Logistic regression models were employed to ascertain the odds ratios (OR) and 95 percent confidence intervals (95% CI) for the association between DII and LTBI. Model 1 was adjusted for social demographic factors, including age,gender,race,education,marital status,smoking,drinking,BMI and PIR. Model 2 was further adjusted for DM and HPT.

Furthermore, restricted cubic spline (RCS) regression was conducted with 4 knots at the 5th, 35th, 65th, and 95th percentiles of DII to evaluate linearity and investigate the dose–response curve between DII and LTBI after adjusting for variables in Model 2.

Moreover, potential alterations in the association between DII and LTBI were evaluated, encompassing the following variables: gender, age (<65vs.≥65 years), BMI (<30 vs. ≥ 30 Kg/m²), PIR (<1.3, 1.3-3.5, ≥3.5),smoking, drinking, HPT and DM.Variability among subgroups was analyzed through multivariate logistic regression, and interactions between subgroups and DII were investigated via likelihood ratio testing.

Since the sample size was established exclusively based on the available data, no prior statistical power assessments were conducted. All analyses were executed utilizing the statistical software packages R 3.3.2 (http://www.R-project.org, The R Foundation, Shanghai, China) (accessed on 26 November 2024) and Free Statistics software version 2.0 [28]. A descriptive study was carried out involving all participants. Through two-tailed testing, a p-value of <0.05 was deemed significant.

## Results

### 1. Study Population

In total, 7153 participants completed the TB test, we excluded those TB data was not determined(n=29), missing data on DII (n=553), and those lacking data on covariates (n=2697). Ultimately, this cross-sectional study comprised 3,892 participants from the NHANES between 2011 and 2012 for the analysis. The detailed inclusion and exclusion process is illustrated in Figure 1.

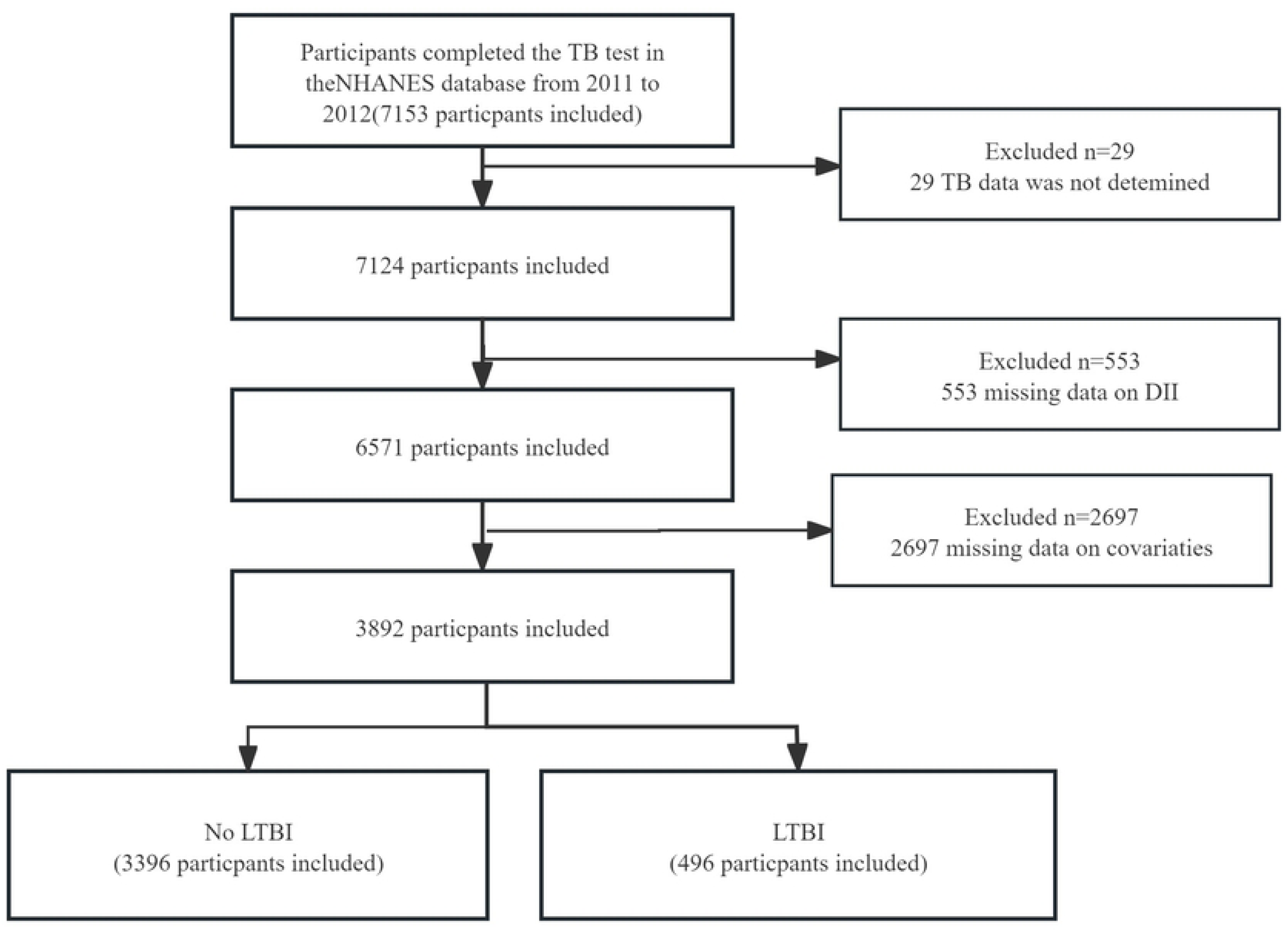

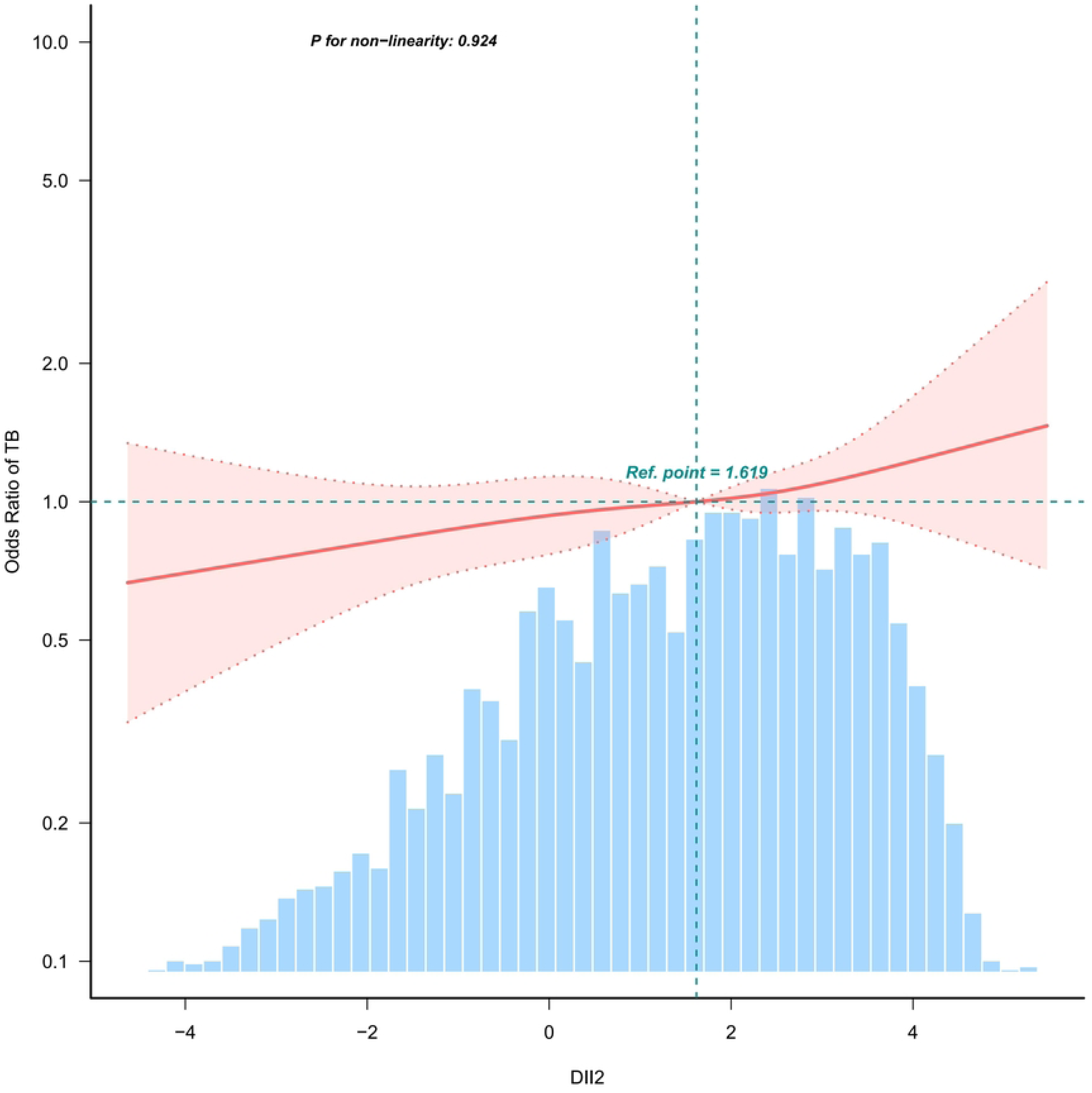

### 2 Baseline Characteristics

A total of 3,923 patients aged 20 to 80 years were incorporated into the analysis.

There were 496 (12.74%) individuals with LTBI. Table 1 presents the general characteristics of the participants based on LTBI. The average age of the participants was 48.4 ±17.8 years, and 1,923 (49.4%) individuals were female. Those participants with a positive LTBI tended to be male,elder, smokers, drinkers,with HPT, DM,Mexican American,other Hispanic or other races,married,possess less than high school educational attainment, and have a lower PIR.

**Table 1.** Population characteristics by categories of LTBI.

| Variables | Total (n = 3892) | LTBI | No-LTBI | *P* value |
| --- | --- | --- | --- | --- |
| **Gender** |  |  |  | < 0.001 |
| Male | 1969 (50.6) | 295 (59.5) | 1674 (49.3) |  |
| Female | 1923 (49.4) | 201 (40.5) | 1722 (50.7) |  |
| **Age** | 48.4 ± 17.8 | 53.5 ± 16.1 | 47.7 ± 17.9 | < 0.001 |
| **BMI** | 29.0 ± 6.9 | 28.8 ± 6.8 | 29.1 ± 7.0 | 0.451 |
| **smoking** |  |  |  | 0.011 |
| Yes | 795 (20.4) | 107 (21.6) | 688 (20.3) |  |
| Former | 910 (23.4) | 139 (28) | 771 (22.7) |  |
| Nerver | 2187 (56.2) | 250 (50.4) | 1937 (57) |  |
| **drinking** |  |  |  | 0.003 |
| No | 2919 (75.0) | 345 (69.6) | 2574 (75.8) |  |
| Yes | 973 (25.0) | 151 (30.4) | 822 (24.2) |  |
| **HPT** |  |  |  | 0.026 |
| Yes | 1410 (36.2) | 202 (40.7) | 1208 (35.6) |  |
| No | 2482 (63.8) | 294 (59.3) | 2188 (64.4) |  |
| **DM** |  |  |  | < 0.001 |
| Yes | 486 (12.5) | 86 (17.3) | 400 (11.8) |  |
| No | 3406 (87.5) | 410 (82.7) | 2996 (88.2) |  |
| **Race** |  |  |  | < 0.001 |
| Mexican American | 358 ( 9.2) | 77 (15.5) | 281 (8.3) |  |
| non-Hispanic White | 1596 (41.0) | 74 (14.9) | 1522 (44.8) |  |
| non-Hispanic Black | 999 (25.7) | 131 (26.4) | 868 (25.6) |  |
| Other Hispanic or other | 939 (24.1) | 214 (43.1) | 725 (21.3) |  |
| **Marital** |  |  |  | 0.002 |
| Married or live with partner | 2181 (56.0) | 310 (62.5) | 1871 (55.1) |  |
| Never married | 1711 (44.0) | 186 (37.5) | 1525 (44.9) |  |
| **Education** |  |  |  | < 0.001 |
| less than high school | 804 (20.7) | 166 (33.5) | 638 (18.8) |  |
| high school or above | 3088 (79.3) | 330 (66.5) | 2758 (81.2) |  |
| **PIR** | 2.0 (1.0, 4.1) | 1.8 (0.9, 3.6) | 2.0 (1.1, 4.2) | 0.004 |
| **DII** | 1.6 (0.0, 2.9) | 1.5 (-0.1, 2.7) | 1.6 (0.1, 3.0) | 0.039 |

### 3 Relationship between DII and LTBI

The univariate analysis demonstrated that gender,age, race, education, marital status,drinking,DM,HPT,PIR,DII were associated with LTBI.However, no significant differences were observed between the groups regarding BMI and smoking (all *P* > 0.05)(Table 2).

**Table 2.**
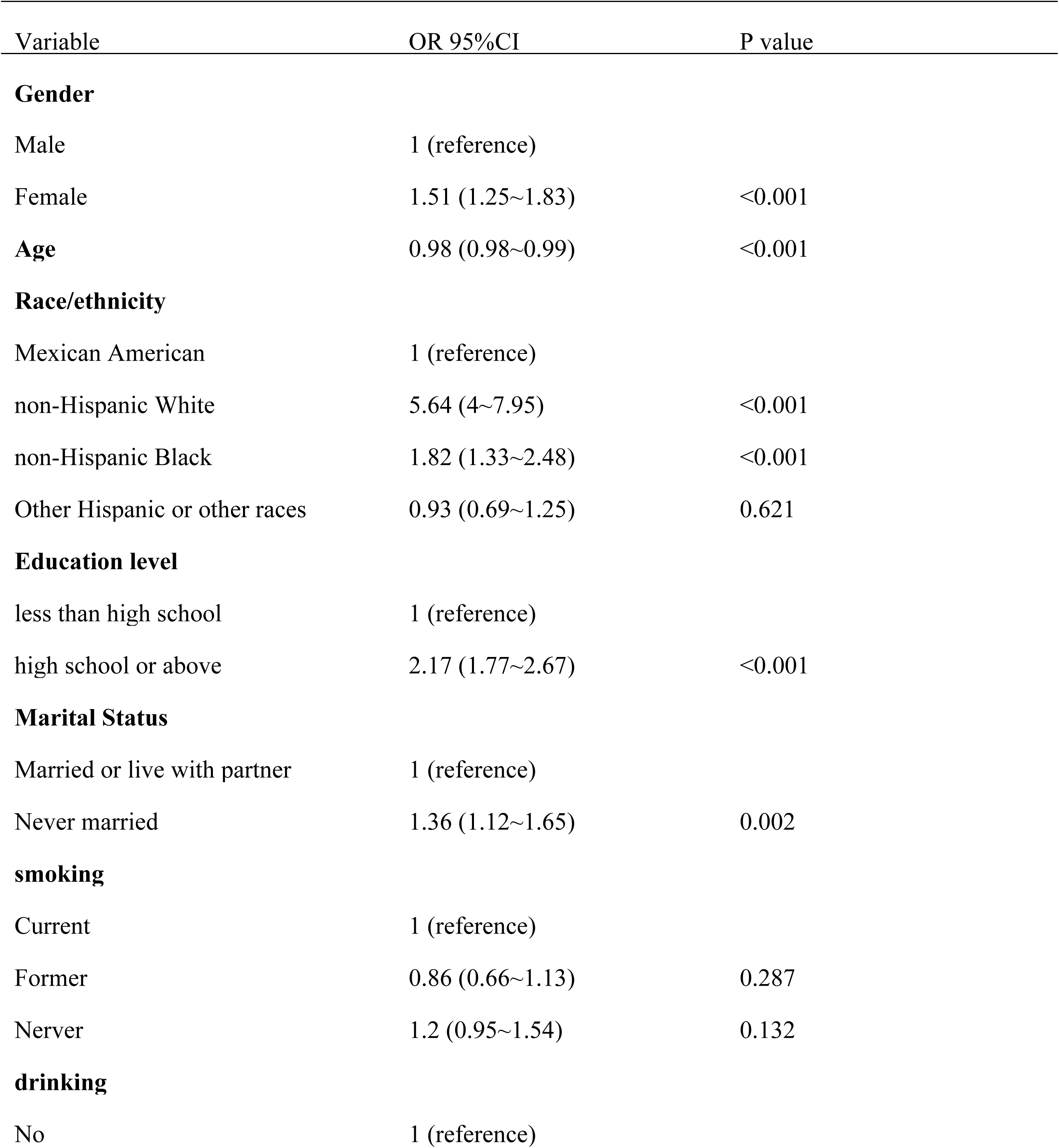
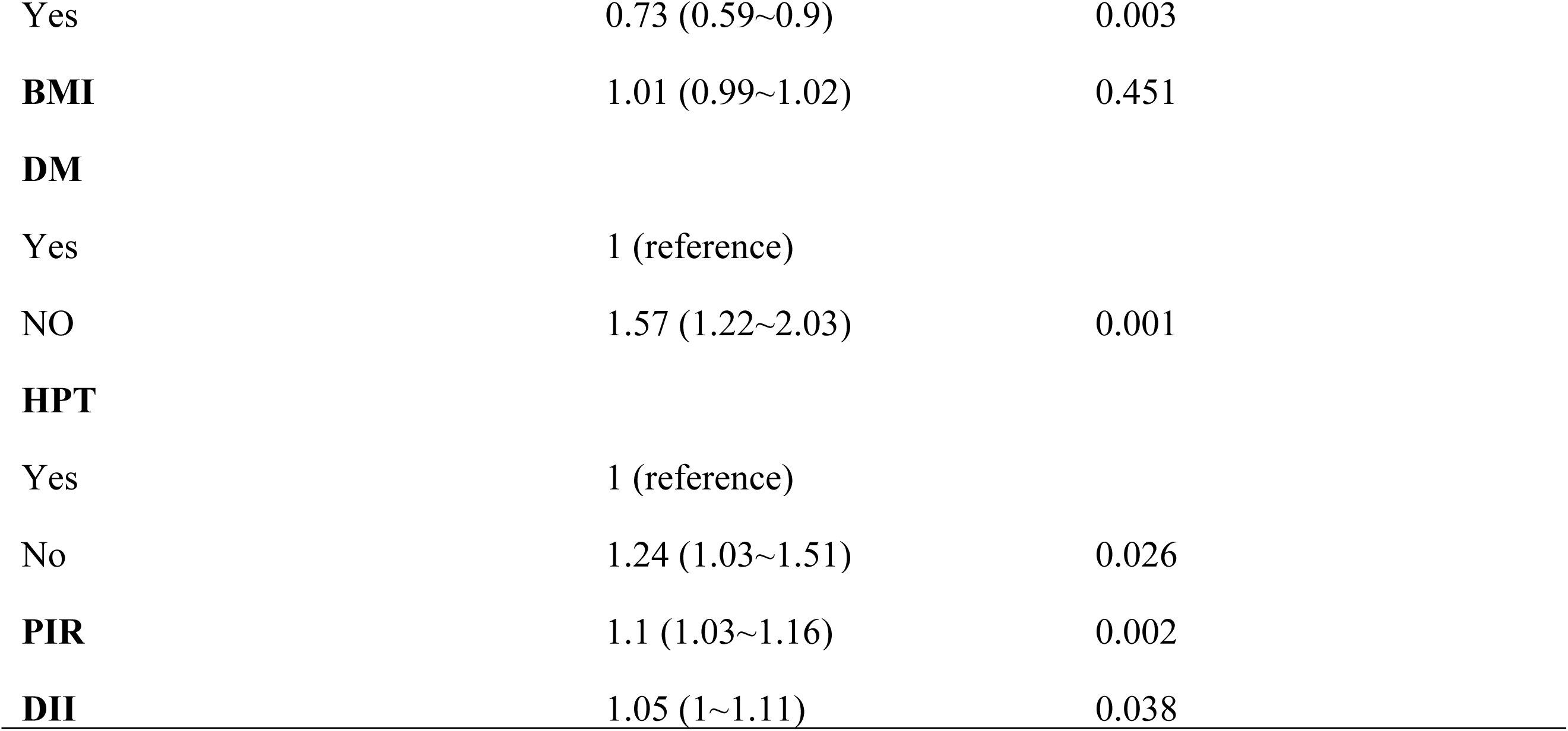
Association of covariates and TB infection.

The findings of the multivariate logistic regression analysis are shown in Table 3.

**Table 3.** Multivariate logistic regression models evaluating the association between DII and LTBI.

| Variable | NO. | OR |  |  |  |  |  |
| --- | --- | --- | --- | --- | --- | --- | --- |
|  |  | Crude | P value | Model 1 | P value | Model 2 | P value |
| DII(continuous) |  |  |  |  |  |  |  |
| DII | 3892 | 1.05 (1~1.11) | 0.038 | 1.06 | 0.025 | 1.07 | 0.023 |
| DII(classified) |  |  |  |  |  |  |  |
| T1 | 1297 | 1(Ref) |  | 1(Ref) |  | 1(Ref) |  |
| T2 | 1297 | 1.07 | 0.566 | 1.07 | 0.563 | 1.08 | 0.548 |
| T3 | 1298 | 1.27 | 0.044 | 1.34 | 0.025 | 1.35 | 0.023 |
| Trend test | 3892 |  | 0.045 |  | 0.026 |  | 0.024 |

In the unadjusted model, DII was shown to be positively related to the risk of LTBI (OR, 1.05; 95% CI, 1.00–1.11). Results were similar after adjusting for age,gender,race,education,marital status,smoking,drinking,BMI and PIR(OR,1.06; 95% CI, 1.01-1.12). After adjusting for other possible confounders, including DM,and HPT, the positive association remained significant (OR,1.07;95% CI, 1.01∼1.13),all *P*-value < 0.05.

Conversion of DII into aliquot 3 categorical variables and performed multiple logistic regression, found that compared with the T1 group,the risk of LTBI was increased by 27% in the T3 group(OR, 1.27 95% CI 1.01 to 1.61), after adjusting for the above covariates, we obtained similar results,model 1(OR,1.34 95%CI 1.04∼1.74),model 2(OR,1.35 95%CI 1.04∼1.75).

Adjusted smoothed plots suggest a straightforward linear relationship between DII and LTBI (Figure 3, P for non-linearity = 0.924, the highest and lowest 0.5% was trimmed for each DII measure.) As the level of DII increases, the risk of LTBI shows an upward trend.

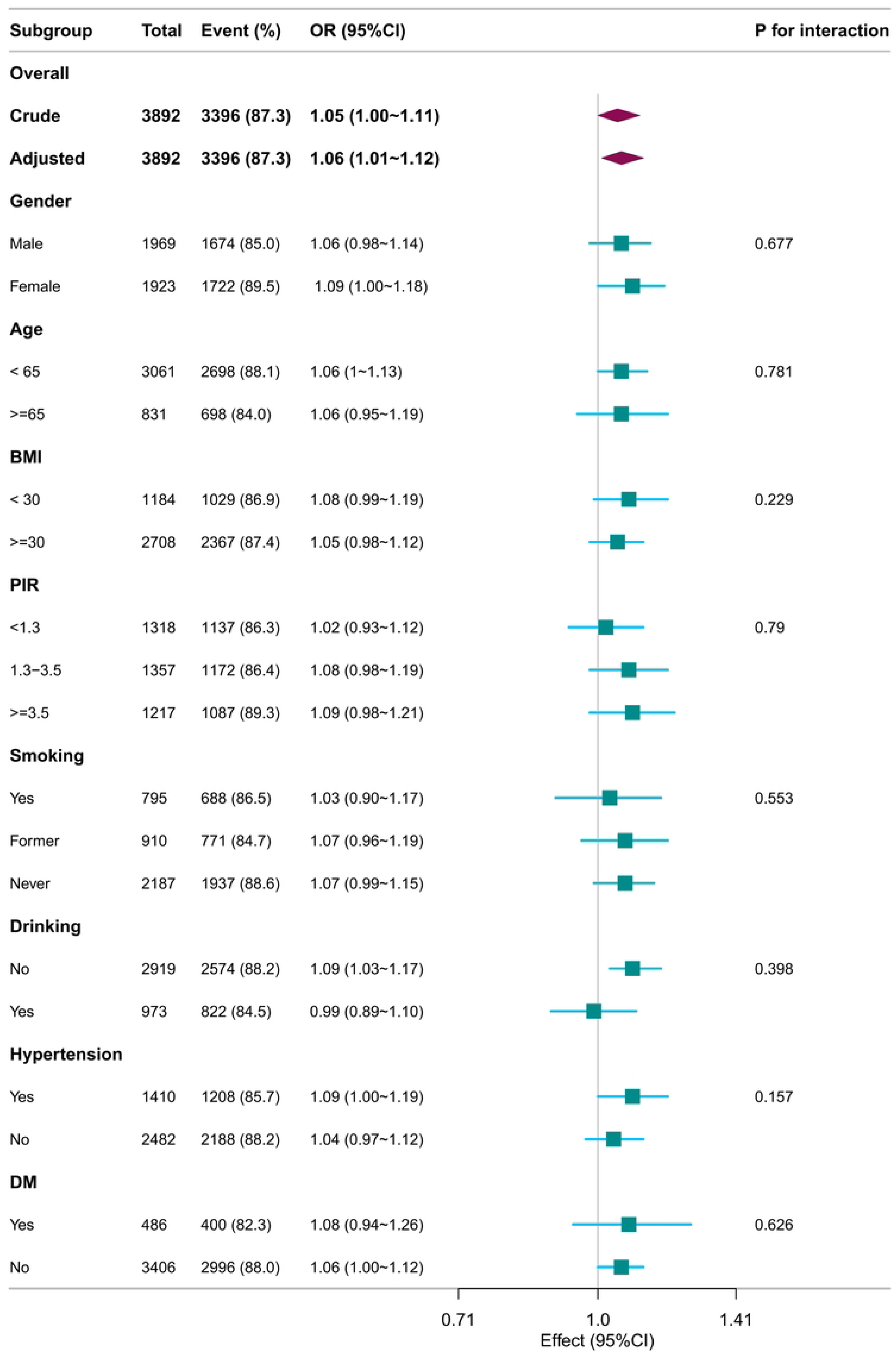

### 4 Stratified Analyses Based on Additional Variables

In several subgroups, stratified analysis was performed to assess potential effect modifications on the relationship between DII and LTBI. No significant interactions were found in any subgroups after stratifying by gender, age,BMI,PIR,smoking,drinking,HPT and DM (Figure 3).

## Discussion

To our knowledge, this is the first report of an association between DII score and LTBI. In this cross-sectional study of 3892 adults, a significant positive association between DII and LTBI was observed, suggesting that greater intake of a pro-inflammatory diet may contribute to the risk of LTBI. This association remained robust after adjusting for the covariates.

The DII has been extensively studied in several observational studies that investigate its association with various health outcomes. For example, research using data from the NHANES found that individuals with higher DII scores were more likely to develop early chronic obstructive pulmonary disease (COPD), suggesting a connection between elevated DII scores and this condition [29].Additionally, higher DII scores are linked to an increased risk of mortality in COPD patients [30].Another important study from the Korean Genome and Epidemiology Study found that individuals in the highest quartile of the energy-adjusted DII (E-DII) had a significantly greater risk of developing gastric diseases, including gastritis and gastric ulcers, during the follow-up period [31]. These findings underscore the DII’s potential as a predictor of chronic diseases and suggest the need for additional research to confirm these associations in diverse populations.

While the current study reveals a significant association between the DII and the risk of LTBI, several limitations must be acknowledged. The cross-sectional design restricts the ability to infer causality, as potential confounding factors may still influence the observed relationship. Additionally, dietary assessments rely on self-reported data, which can introduce bias and inaccuracies due to recall errors or misreporting. The study also lacks longitudinal follow-up, which would provide insight into changes in dietary habits and their long-term effects on LTBI risk.

Furthermore, the generalizability of the findings may be limited to specific populations, necessitating caution when extrapolating to diverse demographic groups.

## Conclusions

In conclusion, our study showed that a pro-inflammatory diet is associated with the risk of developing LTBI. Given that TB infection is a major global public health problem, identifying modifiable risk factors for preventing and treating TB infection from a population medicine perspective is imperative.

## Data Availability

Publicly available datasets are available online for this study. The repository/repositories name and accession numbers are available online at http://www.cdc.gov/nchs/nhanes.htm (accessed on 26 November 2024).

## Acknowledgment

We are grateful to thank all of the participants for their valuable contributions.

We appreciatively thank Dr. Jie Liu (Department of Vascular and Endovascular Surgery, Chinese PLA General Hospital) and Dr. Huanxian Liu (Department of Neurology, Chinese PLA General Hospital)for his consultation on the study design, language polishing, proofreading, statistical support, and comments regarding the manuscript.

## Author Contributions

Author 1(First author): Jingbo Jia collected, analyzed and interpreted the data and results, and drafted the manuscript.

Author 2 (Co-first author): Yuanyuan Liu proposed the concept and design of the study and revised manuscript for critical intellectual content.

Author 3: Hua Zhang performed data cleaning of this paper, guidance on statistical analysis and revision of the manuscript.

Author 4: Jianrui Pi was responsible for implementing the study and collecting the data.

Author 5: Chao Wang is responsible for collecting the data and analyzing the data. Author 6: Shiqi Dong was responsible for implementing the study and collecting the data.

Author 7: Tong Zhang was responsible for implementing the study and drafting the article.

Corresponding author:Wanjie Yang revised manuscript for critical intellectual content.

All authors had access to the data. All authors read and approved the final manuscript.

## Disclosure statement

The authors have no conflicts of interest.

Ethics approval was obtained from the NCHS Ethics Review Committee, and participants provided written informed consent. The secondary analysis did not require additional institutional review board approval.

## Data Availability Statement

Publicly available datasets are available online for this study. The repository/repositories name and accession numbers are available online at http://www.cdc.gov/ nchs/nhanes.htm (accessed on 26 November 2024).

